# Increasing emergency department referral of chest pain patients for non-invasive cardiac testing does not improve two-year clinical outcomes

**DOI:** 10.1101/2023.08.18.23294292

**Authors:** Dustin G. Mark, Jie Huang, Dustin W. Ballard, David R. Vinson, Jamal S. Rana, Dana R. Sax, Adina S. Rauchwerger, Mary E. Reed, the Kaiser Permanente CREST Network Investigators

## Abstract

**Background:** Non-invasive cardiac testing (NICT) has been associated with decreased long-term risks of major adverse cardiac events (MACE) among emergency department (ED) patients at high coronary risk. It is unclear whether this association extends to patients without evidence of myocardial injury on initial electrocardiogram and cardiac troponin testing.

**Methods:** Retrospective cohort study of patients presenting with chest pain between 2013-2019 to 21 EDs within an integrated health care system, excluding patients with ST-elevation myocardial infarction or myocardial injury by serum troponin testing. To account for confounding by indication, we grouped patient encounters by the NICT referral rate of the initially assigned emergency physician, relative to local peers within discrete time periods. The primary outcome was MACE within two years. Secondary outcomes were coronary revascularization and MACE inclusive of all-cause mortality (MACE-ALL). Associations between NICT referral group (low, intermediate, or high) and outcomes were assessed using risk-adjusted proportional hazards methods with censoring for competing events.

**Results:** Among 144,577 eligible patient encounters, 30-day NICT referral was 13.0%, 19.9% and 27.8% in low, intermediate, and high NICT referral groups, respectively, with good balance of baseline covariates between groups. Compared with the low group, there was no significant decrease in the adjusted hazard ratio (aHR) of MACE within the intermediate (aHR 1.08, 95% CI 1.02-1.14, adjusted p = 0.024) or high (aHR 1.05, 95% CI 0.99-1.11, adjusted p = 0.13) NICT referral groups. Results were similar for MACE-ALL and coronary revascularization, as well as subgroup analyses stratified by estimated risk (HEART score; 48.2% low-risk, 49.2% moderate-risk, 2.7% high-risk).

**Conclusion:** Increased NICT referral was not associated with a decreased hazard of MACE within two years following ED visits for chest pain without evidence of acute myocardial injury. These findings further highlight the need for evidence-based guidance regarding appropriate use of NICT in this population.

**What is Known:**

- In emergency department patients with chest pain but no evidence of acute myocardial injury, referral for non-invasive cardiac testing is not associated with improved outcomes within 6 to 12 months.
- There is some evidence that non-invasive cardiac testing may improve longer-term outcomes, especially among patients at moderate to high risk of coronary disease.

**What the Study Adds:**

- Higher non-invasive cardiac testing was not associated with improved outcomes within two years following emergency department visits for chest pain, absent evidence of acute myocardial injury.
- This finding was most evident among patients at low to moderate predicted coronary risk.

## Introduction

Chest pain is a leading reason for emergency department (ED) visits in the U.S., with upwards of 7 million patients evaluated annually.^1^ Prior to 2021, American Heart Association/American College of Cardiology guidelines recommended functional or anatomic non-invasive cardiac testing (NICT) of ED patients with chest pain and possible acute coronary syndrome, irrespective of estimated risk, prior to or within 72 hours of ED discharge.^2^ However, several large observational studies of ED patients with follow-up periods ranging from 30 to 365 days have noted that NICT was only associated with improved outcomes among patients with high estimated coronary risk and/or elevated serum cardiac troponin values.^3–12^ Accordingly, newer guidelines and clinical policies have recommended against the routine use of NICT for ED patients at low estimated risk of acute coronary syndrome.^13–15^

However, it remains possible that there are longer-term benefits of NICT for patients at low or intermediate risk, in part because NICT does appear to increase the likelihood of coronary revascularization in these populations.^4,12,16^ The SCOT-HEART randomized clinical trial of CT coronary angiography vs. standard care among low-to intermediate-risk patients with stable chest pain demonstrated a reduction in rates of non-fatal myocardial infarction beginning approximately 1.5 years of follow-up, an effect that appeared driven by preventative medical therapies and/or earlier coronary revascularizations.^17^ A propensity-matched study of ED patients who underwent NICT within 30 days after a chest pain evaluation showed a non-significant reduction in the composite outcome of myocardial infarction or cardiovascular mortality at 1 year among the intermediate-risk group (HR 0.88, 95% CI 0.77-1.01).^5^ Finally, an observational study of roughly 1.5 million patients in the outpatient Canadian setting examined NICT completion within 90 days of a visit for chest pain or angina and found a 25% reduction in a composite outcome of unstable angina, myocardial infarction, and cardiovascular mortality over a median of four years of follow-up, with statistically significant reductions in each of the individual outcome constituents.^18^ Thus, it is conceivable that risk reduction among lower-risk patients is more of an incremental and gradual process than among high-risk patients.

To help further examine this question, we previously utilized the intrinsic variability in NICT utilization that exists at both the physician and medical center level to examine associations between NICT and outcomes in a quasi-experimental observational fashion by creating well-balanced groups of ED patients with low, intermediate, or high referrals for NICT. ^8,10,16,19,20^ In that study, we did not observe an association between higher NICT utilization and 60-day major adverse cardiac events (MACE), though we did observe a positive association between NICT and coronary revascularization, consistent with other studies.^4,12^ We now report two-year outcomes from that study population, with additional differentiation between cardiac and all-cause mortality. Our hypothesis was that higher NICT utilization would be associated with decreased risk of two-year MACE, particularly among patients at intermediate to high risk for coronary disease.

## Methods

### Study Design and Setting

We performed a retrospective study of ED chest pain encounters between January 1, 2013, and December 31, 2019, across 21 community EDs within Kaiser Permanente Northern California (KPNC). KPNC is a private, non-profit integrated healthcare system that covers 4.3 million members representing one-third of the region’s population. KPNC members have been found to be comparable to the surrounding population with respect to age, sex, race and ethnicity.^21,22^ All care facilities (emergency, outpatient, inpatient) within KPNC utilize the same comprehensive integrated electronic health record (EHR; Epic, Verona, WI). In this study setting, patients are randomly assigned to emergency physicians in a rotating fashion upon intake and triage. The study was approved by the KPNC Institutional Review Board with a waiver of informed consent.

### Patient Selection

We included patient encounters with a chief complaint or ED diagnosis of chest pain as well as a serum troponin level measurement in the ED. To capture outcomes in a consistent manner, we required active membership in the health system for two years following the index visit, except in cases of death. We excluded patients aged under 30 or over 80 years (to select for patients most likely to benefit from NICT) or if there was a cardiac troponin value above the 99^th^ percentile upper reference limit during the ED visit, an index ED diagnosis of ST-elevated myocardial infarction (as initial cardiac troponin levels may be normal in these patients^23^), an ED disposition of “expired” or “left against medical advice”, an objective cardiac test performed within the prior 6 months (exercise treadmill, myocardial perfusion, stress echo, computed tomography [CT] coronary angiography, cardiac catheterization), or if the assigned ED physician had less than 100 eligible encounters for a given epoch (inadequate sampling for NICT referral ranking, as defined below). In cases of multiple encounters by the same patient during the study period, only the first study eligible ED encounter within a given two-year window was included to avoid overlapping outcome periods.

### Baseline Variables

For each patient encounter, study variables were obtained from the EHR using automated electronic data extraction. This methodology was previously validated using both manually abstracted and prospectively collected data, with similar levels of agreement as compared to other prospective interrater data validation studies among ED chest pain patients.^24,25^ Variables included age, past medical history, peak troponin value in the ED, and both presenting symptoms and electrocardiogram (ECG) findings using text string analysis of clinical notes and final ECG interpretations, respectively (Methods Supplement). We retrospectively calculated a history, electrocardiogram, age, risk factors, troponin (HEART) score for each eligible patient encounter (intraclass correlation coefficient of 0.84 as compared to prospective data).^25^ Since patients with troponin values above the 99^th^ percentile were excluded from the study, the maximum HEART score was 8 as opposed to 10 in the original HEART score validation study.^26^ For the history component of the HEART score (range 0-2 points), we weighted higher risk symptoms (e.g., chest pressure or radiation to the arms or shoulders) against lower risk symptoms (e.g., pain worse with inspiration or pain reproduced with palpation) in a standardized fashion to arrive at a net balance of risk represented by the patient’s symptoms.^27^

Troponin values were obtained using the Access AccuTnI assay from 2013 through July 14, 2014 and the Access AccuTnI+3 assay from July 15, 2014 through 2019 (both Beckman-Coulter, Brea, CA). The 99^th^ percentile upper reference limit for both assays is 0.04 ng/mL per local institutional reporting guidelines and reference literature.^28^ Because high-normal range troponins have prognostic value independent of the HEART score, we treated absolute troponin value an independent risk factor categorized as either below the limit of quantitation (LOQ, <0.02 ng/ml) or within the quantifiable normal range (between 0.02-0.04 ng/ml).^24^

### NICT referral groups

As described previously^16^, to help account for confounding by indication, we utilized emergency physician-level variation in NICT referral. This approach takes advantage of the significant variance among emergency physicians in their tendency to refer for NICT and the context of a random patient-to-physician assignment system used within the study setting. We ranked emergency physicians based on how often their study-eligible patients were referred for NICT within 72 hours of the ED encounter. This ranking was relative to other emergency physicians in the local practice (to control for interfacility practice variation) and was repeated during three discrete 28-month epochs of time covering the entire study period (to account for personnel changes within local practices). To improve ranking accuracy, we excluded encounters if the emergency physician had less than 100 study-eligible patient encounters within a given epoch. Encounters were then categorized into one of three groups of NICT referral (low, intermediate, high) based on the facility and epoch-specific relative ranking of the initially assigned emergency physician. The distribution of baseline covariates between NICT referral groups was assessed using standardized differences with a value less than 0.10 indicating acceptable covariate balance.^29^

### Outcomes

The primary outcome was MACE within a two-year follow-up period from the index encounter. MACE was defined as the composite of myocardial infarction, cardiac arrest, cardiogenic shock, or cardiac death. Myocardial infarction, cardiac arrest, and cardiogenic shock were defined based on the presence of corresponding International Classification of Disease, 9^th^ and 10^th^ revision (ICD-9/10) diagnosis codes (ICD-9 for January 1, 2013 through September 30, 2015; ICD-10 for October 1, 2015 through December 31, 2019) listed as the first or second diagnosis at an inpatient or ED encounter within the integrated health care system, or in a coded claim for services provided at a facility outside the integrated health care system. We determined mortality using a composite death database drawing from KPNC mortality records, California Department of Public Health Vital Records, the Social Security Death Index, and the National Death Index (for cause of death). Cardiac death was designated if an ICD-10 code for “diseases of the heart” (as per the National Vital Statistics System) was listed as an underlying cause of death.^30^ Secondary outcomes were coronary revascularization (percutaneous or surgical) and MACE inclusive of all-cause mortality (MACE-ALL), both within a two-year follow-up period. Coronary revascularization was deemed to have occurred if any corresponding ICD-9/10 procedure or current procedural technology (CPT) code was used during a hospitalization within or outside the integrated health care system. Coronary revascularization was treated as a separate outcome (as opposed to being included within the composite MACE definition) because there is no reliable method to differentiate between elective or non-elective coronary revascularizations using diagnostic and/or billing codes, and elective coronary revascularization procedures are inconsistent with consensus agreements on appropriate composite MACE definitions. ^31,32^ ICD-9/10 and CPT codes used for the outcome definitions above are detailed in Methods Supplement 2. To describe downstream processes of care, we measured the following within 30 days of an encounter: performance of objective cardiac testing (NICT or cardiac catheterization), new prescriptions for cardiovascular medications (no prior fills from that drug class in the 120 days prior) and outpatient visits with primary care physicians or cardiologists.

### Statistical Analysis

Associations between the rate of MACE, MACE-ALL or coronary revascularization and NICT referral group were assessed using mixed-effects proportional hazard models adjusted for patient age, sex, individual risk factors, peak troponin in the ED, ischemic-appearing ECG, and study epoch, with random effects at the facility level and robust standard errors. For MACE and coronary revascularization, we determined the cause-specific hazard by censoring for competing outcomes (non-cardiac death for MACE and all-cause mortality for coronary revascularization).^33^ The low referral group served as the reference category. These models were applied both to the overall study population and separately to patients within the three strata of HEART score (low-risk = 0-3; moderate-risk = 4-6; high-risk = > 6). An α=0.05 was used to define statistical significance of hypothesis tests, with p-values adjusted for multiple comparisons using the Holm-Bonferroni method.^34^ Data analyses were performed using Stata 17.0 (StataCorp, TX).

### Sensitivity Analysis

Since previous findings showed a general decline in the completion of objective cardiac testing within 30 days of the index ED visit during the third study epoch^35^, and to account for potential impact of the COVID-19 pandemic on two-year follow-up period^36^, we repeated the overall statistical analysis using encounters from only the first and second study epochs (January 1, 2013 through August 31, 2017). We also repeated the primary analysis among a restricted subgroup of encounters with troponin values in the quantifiable range of normal (0.02 to 0.04 ng/ml) due to the higher independent risk of MACE in these patients, as noted above. Given that proportional hazards do not indicate the magnitude of association between a covariate and a dependent variable^37^, we alternatively examined the association between NICT referral group and two-year MACE-ALL using mixed-effects logistic regression. Finally, as a post-hoc exploratory analysis, we assessed for interactions between the high and low NICT referral groups and several covariates (age, known coronary artery disease, HEART risk score category, and peak troponin value).

## Results

There were 144,577 included ED encounters (Figure 1) with 700 emergency physicians across the 7-year study period. HEART score distribution was 48.2% low-risk, 49.2% moderate-risk and 2.7% high-risk. The median age was 58 years (interquartile range 48-68 years), 57% were female, 23% had diabetes, 13% had coronary artery disease, 3.9% had ischemic ECG findings, 12% had a peak troponin between the LOQ and the 99^th^ percentile, and 20% were referred for NICT within 72 hours of the ED encounter (Table 1). Baseline demographic and risk factors were well-balanced across NICT referral groups, and the frequency of NICT referral was significantly different between low, intermediate, and high groups (13.0%, 19.9%, and 27.8%, p<0.0001, Table 1). Covariate balance was maintained within HEART score risk strata with similar between-group differences in NICT referral (Table S1).

**Figure 1.**
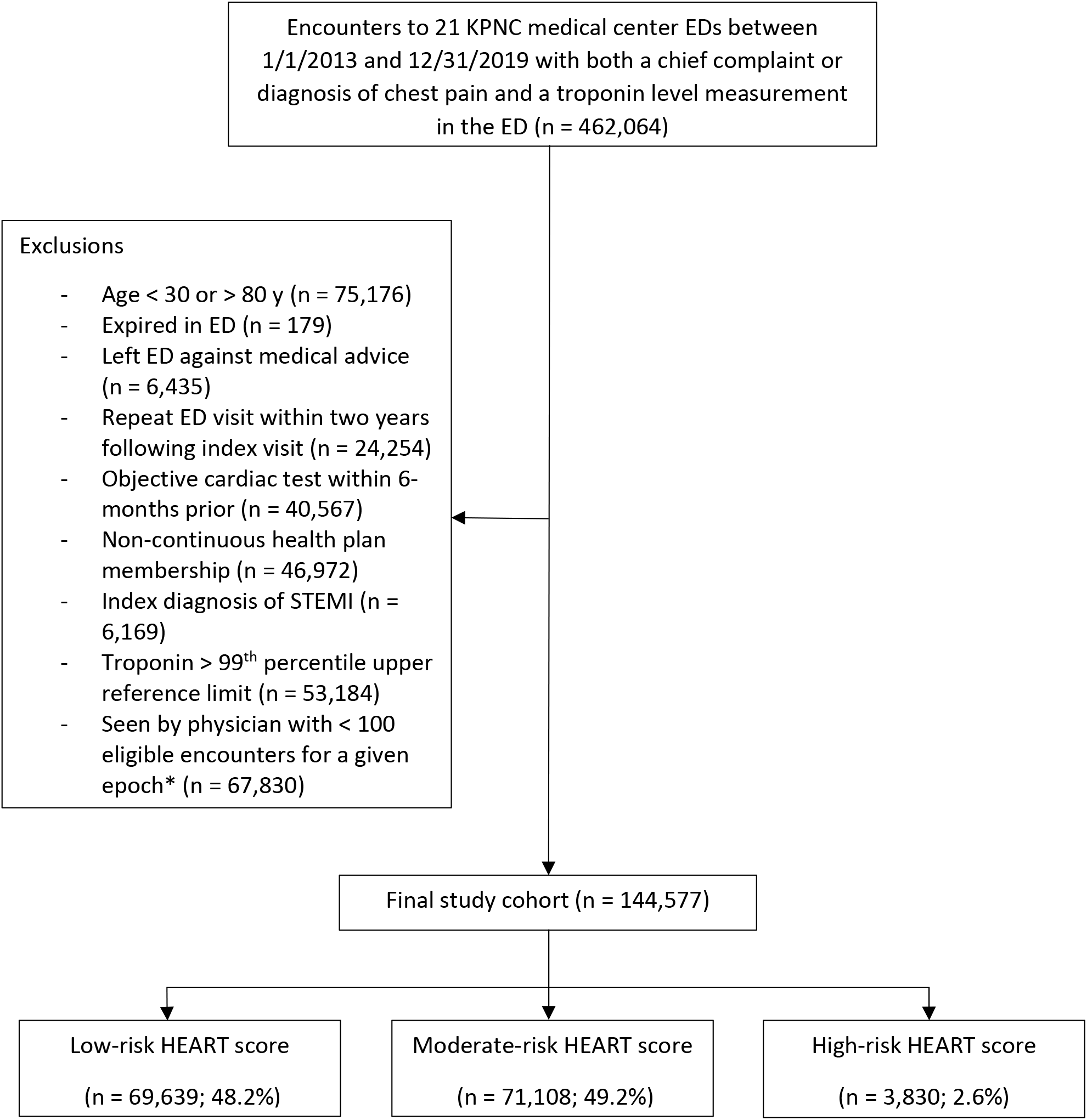
Cohort selection. *To account for practice pattern changes due to time as a confounder in our ranking of ED provider NICT referral-intensity, we divided the 7-year study period into three equal (28-month) epochs: 1) January 1, 2013 through April 30, 2015; 2) May 1, 2015 through August 31, 2017; 3) September 1, 2017 through December 31, 2019. Abbreviations: ED = emergency xsxsdepartment; HEART = history, electrocardiogram, age, risk factors, troponin; KPNC = Kaiser Permanente Northern California; STEMI = ST-elevated myocardial infarction.

**Table 1.**
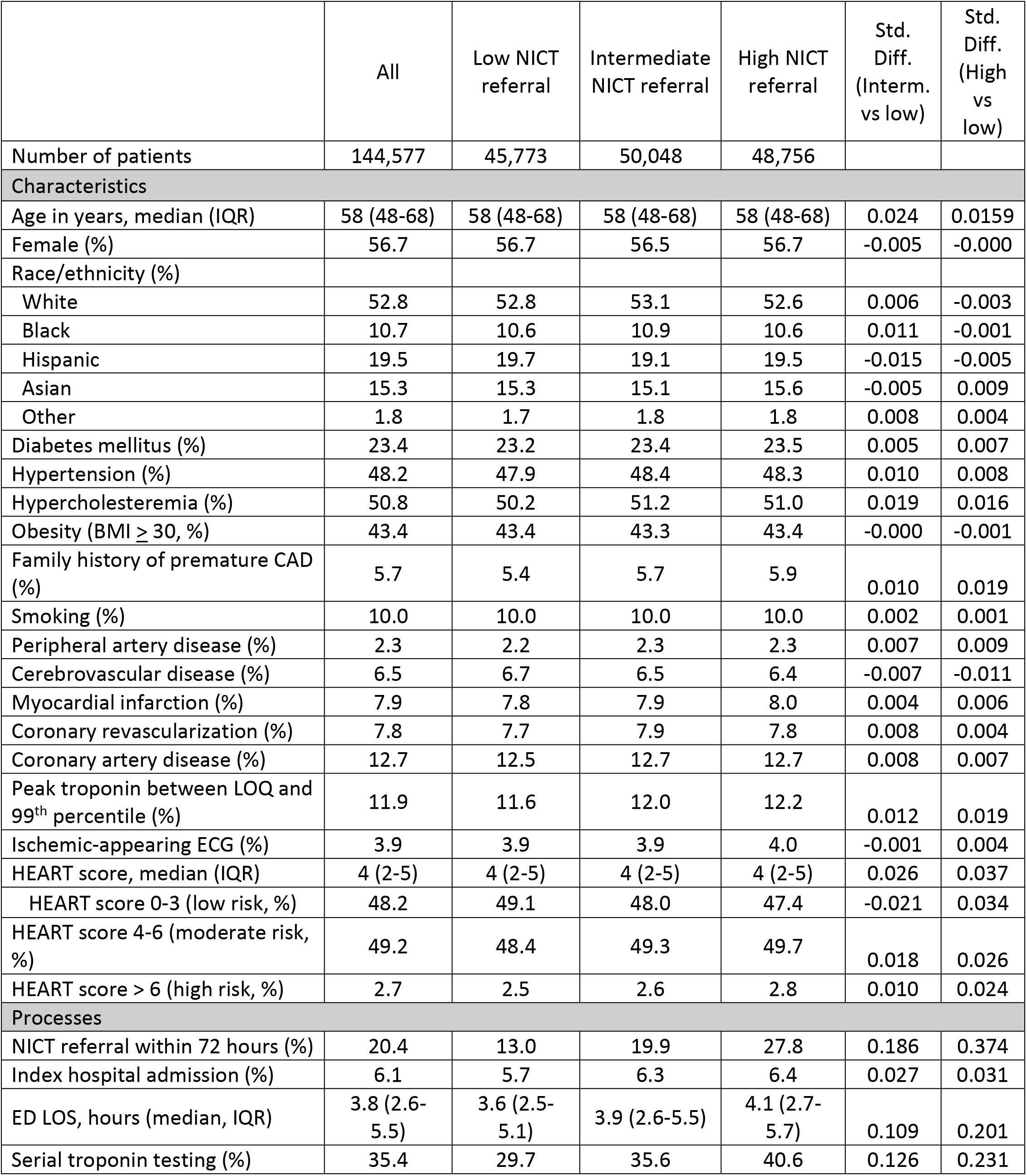
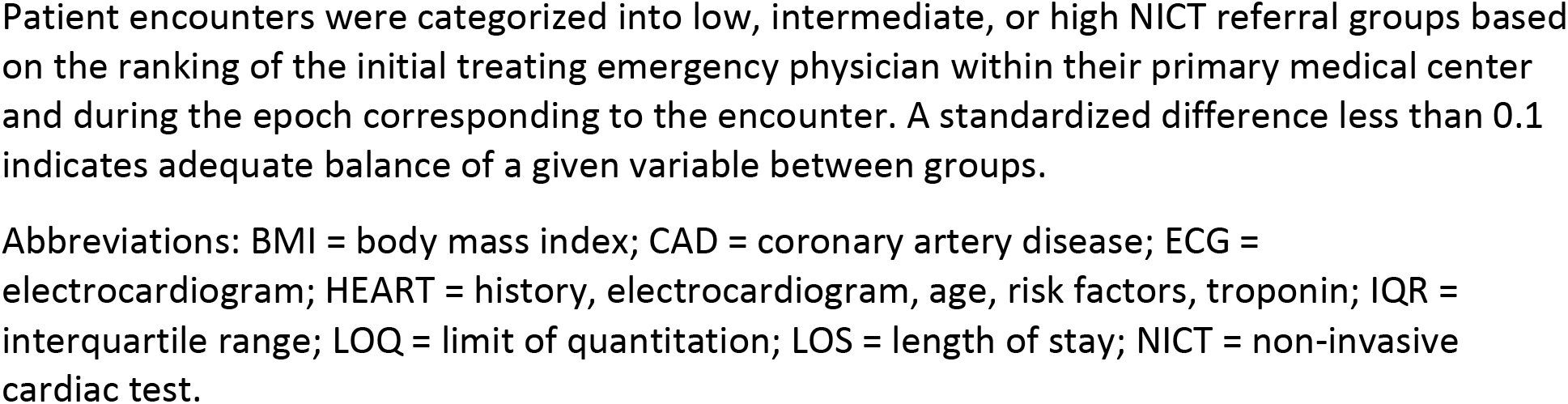
Cohort characteristics.

|  | All | Low NICT referral | Intermediate NICT referral | High NICT referral | Std. Diff. (Interm. vs low) | Std. Diff. (High vs low) |
| --- | --- | --- | --- | --- | --- | --- |
| Number of patients | 144,577 | 45,773 | 50,048 | 48,756 |  |  |
| **Characteristics** |  |  |  |  |  |  |
| Age in years, median (IQR) | 58 (48-68) | 58 (48-68) | 58 (48-68) | 58 (48-68) | 0.024 | 0.0159 |
| Female (%) | 56.7 | 56.7 | 56.5 | 56.7 | -0.005 | -0.000 |
| Race/ethnicity (%) |  |  |  |  |  |  |
| White | 52.8 | 52.8 | 53.1 | 52.6 | 0.006 | -0.003 |
| Black | 10.7 | 10.6 | 10.9 | 10.6 | 0.011 | -0.001 |
| Hispanic | 19.5 | 19.7 | 19.1 | 19.5 | -0.015 | -0.005 |
| Asian | 15.3 | 15.3 | 15.1 | 15.6 | -0.005 | 0.009 |
| Other | 1.8 | 1.7 | 1.8 | 1.8 | 0.008 | 0.004 |
| Diabetes mellitus (%) | 23.4 | 23.2 | 23.4 | 23.5 | 0.005 | 0.007 |
| Hypertension (%) | 48.2 | 47.9 | 48.4 | 48.3 | 0.010 | 0.008 |
| Hypercholesteremia (%) | 50.8 | 50.2 | 51.2 | 51.0 | 0.019 | 0.016 |
| Obesity (BMI ≥ 30, %) | 43.4 | 43.4 | 43.3 | 43.4 | -0.000 | -0.001 |
| Family history of premature CAD (%) | 5.7 | 5.4 | 5.7 | 5.9 | 0.010 | 0.019 |
| Smoking (%) | 10.0 | 10.0 | 10.0 | 10.0 | 0.002 | 0.001 |
| Peripheral artery disease (%) | 2.3 | 2.2 | 2.3 | 2.3 | 0.007 | 0.009 |
| Cerebrovascular disease (%) | 6.5 | 6.7 | 6.5 | 6.4 | -0.007 | -0.011 |
| Myocardial infarction (%) | 7.9 | 7.8 | 7.9 | 8.0 | 0.004 | 0.006 |
| Coronary revascularization (%) | 7.8 | 7.7 | 7.9 | 7.8 | 0.008 | 0.004 |
| Coronary artery disease (%) | 12.7 | 12.5 | 12.7 | 12.7 | 0.008 | 0.007 |
| Peak troponin between LOQ and 99<sup>th</sup> percentile (%) | 11.9 | 11.6 | 12.0 | 12.2 | 0.012 | 0.019 |
| Ischemic-appearing ECG (%) | 3.9 | 3.9 | 3.9 | 4.0 | -0.001 | 0.004 |
| HEART score, median (IQR) | 4 (2-5) | 4 (2-5) | 4 (2-5) | 4 (2-5) | 0.026 | 0.037 |
| HEART score 0-3 (low risk, %) | 48.2 | 49.1 | 48.0 | 47.4 | -0.021 | 0.034 |
| HEART score 4-6 (moderate risk, %) | 49.2 | 48.4 | 49.3 | 49.7 | 0.018 | 0.026 |
| HEART score > 6 (high risk, %) | 2.7 | 2.5 | 2.6 | 2.8 | 0.010 | 0.024 |
| **Processes** |  |  |  |  |  |  |
| NICT referral within 72 hours (%) | 20.4 | 13.0 | 19.9 | 27.8 | 0.186 | 0.374 |
| Index hospital admission (%) | 6.1 | 5.7 | 6.3 | 6.4 | 0.027 | 0.031 |
| ED LOS, hours (median, IQR) | 3.8 (2.6-5.5) | 3.6 (2.5-5.1) | 3.9 (2.6-5.5) | 4.1 (2.7-5.7) | 0.109 | 0.201 |
| Serial troponin testing (%) | 35.4 | 29.7 | 35.6 | 40.6 | 0.126 | 0.231 |

Patient encounters were categorized into low, intermediate, or high NICT referral groups based on the ranking of the initial treating emergency physician within their primary medical center and during the epoch corresponding to the encounter. A standardized difference less than 0.1 indicates adequate balance of a given variable between groups.
Abbreviations: BMI = body mass index; CAD = coronary artery disease; ECG = electrocardiogram; HEART = history, electrocardiogram, age, risk factors, troponin; IQR = interquartile range; LOQ = limit of quantitation; LOS = length of stay; NICT = non-invasive cardiac test.

Unadjusted downstream 30-day processes of care and two-year outcomes are presented in Table 2. Higher NICT referral was associated with higher 30-day objective cardiac testing (53.0% versus 32.8% in high versus low groups) along with small increases in new cardiovascular medication prescriptions and outpatient visits with either primary care or cardiology. Overall observed MACE and MACE-ALL within two years was 4.8% and 7.7%, respectively. On adjusted survival analyses (Table 3), neither intermediate or high NICT referral were associated with a lower adjusted hazard ratio (aHR) for MACE (aHRs of 1.08 and 1.05, respectively) or MACE-ALL (aHRs of 1.06 and 1.04, respectively), with lower bounded 95% confidence intervals between 0.99 and 1.02. Analysis by HEART score risk strata revealed similar differences between NICT groups in 30-day objective cardiac testing (Table S2), but likewise no evidence of lower adjusted hazards of MACE or MACE-ALL between NICT referral groups within any of the three HEART score risk strata (Table 4). There were likewise no statistically significant differences in coronary revascularizations in any of the above analyses.

**Table 2.** Unadjusted 2-year outcomes and 30-day cardiac testing by non-invasive cardiac test (NICT) referral group.

|  | All | Low NICT referral | Intermediate NICT referral | High NICT referral | p-value |
| --- | --- | --- | --- | --- | --- |
| Number of patients | 144,577 | 45,773 | 50,048 | 48,756 |  |
| **2-year outcomes (%)** |  |  |  |  |  |
| Acute myocardial infarction | 3.4 | 3.2 | 3.6 | 3.5 | 0.0042 |
| Cardiac arrest | 0.5 | 0.6 | 0.5 | 0.5 | 0.5070 |
| Cardiogenic shock | 0.2 | 0.2 | 0.3 | 0.2 | 0.2663 |
| Coronary revascularization | 2.8 | 2.7 | 2.8 | 2.9 | 0.1395 |
| Cardiac mortality | 1.3 | 1.3 | 1.3 | 1.2 | 0.2539 |
| All-cause mortality | 4.6 | 4.5 | 4.7 | 4.5 | 0.2375 |
| MACE | 4.8 | 4.6 | 5.0 | 4.8 | 0.0102 |
| MACE-ALL | 7.7 | 7.4 | 7.9 | 7.8 | 0.0148 |
| **30-day mediators (%)** |  |  |  |  |  |
| Objective cardiac testing | 43.2 | 32.8 | 43.0 | 53.0 | <.0001 |
| Exercise treadmill | 34.2 | 25.9 | 34.1 | 42.1 | <.0001 |
| Myocardial perfusion | 11.3 | 8.1 | 11.3 | 14.3 | <.0001 |
| Stress echocardiography | 0.7 | 0.6 | 0.7 | 0.9 | 0.0005 |
| CT coronary angiography | 0.5 | 0.4 | 0.5 | 0.6 | 0.0005 |
| Cardiac catheterization | 3.9 | 3.6 | 3.8 | 4.1 | <.0001 |
| Outpatient follow-up | 55.3 | 53.6 | 55.5 | 56.7 | <.0001 |
| Primary care | 52.6 | 51.1 | 52.8 | 53.8 | <.0001 |
| Cardiology | 7.3 | 6.7 | 7.3 | 7.8 | <.0001 |
| New cardiovascular medication prescription | 10.3 | 9.6 | 10.3 | 10.8 | <.0001 |
| ACE inhibitor or ARB | 3.1 | 3.1 | 3.1 | 3.2 | 0.2014 |
| Antiplatelet | 2.8 | 2.5 | 2.7 | 3.3 | <.0001 |
| Beta-blocker | 4.4 | 4.1 | 4.6 | 4.6 | 0.0002 |
| Statin | 3.5 | 3.4 | 3.5 | 3.6 | 0.1845 |

Abbreviations: ACE = angiotensin converting enzyme; ARB = angiotensin receptor blocker; CT = computed tomography; ECG = electrocardiography; MACE = major adverse cardiac event; MACE-ALL = major adverse cardiac event, including all-cause mortality; NICT = non-invasive cardiac test.

**Table 3.** Adjusted hazards of primary and secondary outcomes according to non-invasive cardiac test (NICT) referral group.

| Outcome | NICT referral group | Adjusted hazard ratio (95% CI) | P-value | Adjusted p-value* |
| --- | --- | --- | --- | --- |
| 2-year MACE | Low | 1.0 (ref) | n/a | n/a |
|  | Intermediate | 1.08 (1.02-1.14) | 0.012 | 0.024 |
|  | High | 1.05 (0.99-1.11) | 0.13 | 0.13 |
| 2-year MACE-ALL | Low | 1.0 (ref) | n/a | n/a |
|  | Intermediate | 1.06 (1.01-1.10) | 0.023 | 0.046 |
|  | High | 1.04 (0.99-1.09) | 0.13 | 0.13 |
| 2-year coronary revascularization | Low | 1.0 (ref) | n/a | n/a |
|  | Intermediate | 1.03 (0.96-1.11) | 0.43 | 0.43 |
|  | High | 1.07 (0.99-1.15) | 0.10 | 0.20 |

Associations between outcomes and NICT referral group were assessed using mixed-effects proportional hazard models adjusting for patient age, sex, risk factors (diabetes, coronary artery disease, coronary revascularization, smoking, hypertension, hypercholesteremia, obesity, family history of premature coronary artery disease), peak troponin value while the emergency department, ischemic-appearing changes on electrocardiogram, and epoch, with censoring of competing outcomes (non-cardiac death for MACE, all-cause mortality for coronary revascularization), random effects at the facility level, and robust standard errors.
\* Holm-Bonferroni method
Abbreviations: MACE = major adverse cardiac event; MACE-ALL = major adverse cardiac event, including all-cause mortality; NICT = non-invasive cardiac test.

**Table 4.**
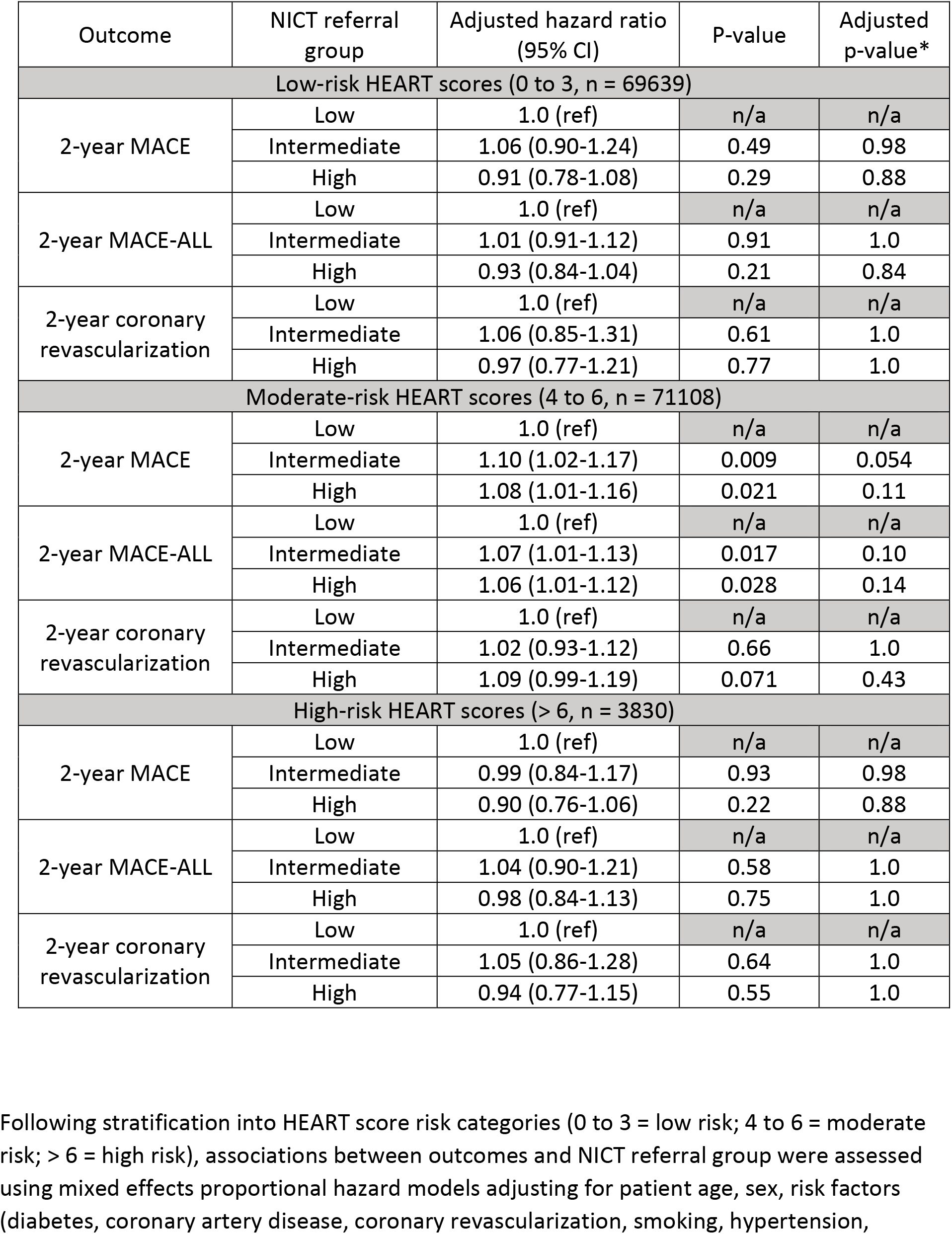
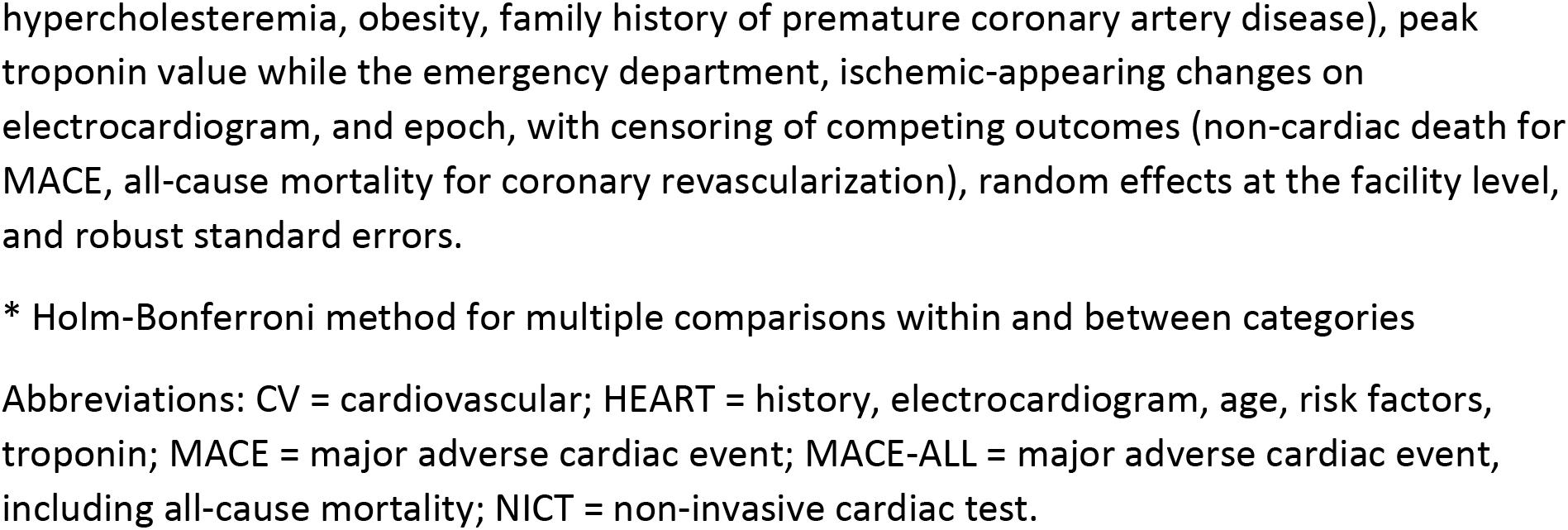
Adjusted primary and secondary outcomes by non-invasive cardiac test (NICT) referral group, stratified by HEART score.

| Outcome | NICT referral group | Adjusted hazard ratio (95% CI) | P-value | Adjusted p-value* |
| --- | --- | --- | --- | --- |
| Low-risk HEART scores (0 to 3, n = 69639) |  |  |  |  |
| 2-year MACE | Low | 1.0 (ref) | n/a | n/a |
|  | Intermediate | 1.06 (0.90-1.24) | 0.49 | 0.98 |
|  | High | 0.91 (0.78-1.08) | 0.29 | 0.88 |
| 2-year MACE-ALL | Low | 1.0 (ref) | n/a | n/a |
|  | Intermediate | 1.01 (0.91-1.12) | 0.91 | 1.0 |
|  | High | 0.93 (0.84-1.04) | 0.21 | 0.84 |
| 2-year coronary revascularization | Low | 1.0 (ref) | n/a | n/a |
|  | Intermediate | 1.06 (0.85-1.31) | 0.61 | 1.0 |
|  | High | 0.97 (0.77-1.21) | 0.77 | 1.0 |
| Moderate-risk HEART scores (4 to 6, n = 71108) |  |  |  |  |
| 2-year MACE | Low | 1.0 (ref) | n/a | n/a |
|  | Intermediate | 1.10 (1.02-1.17) | 0.009 | 0.054 |
|  | High | 1.08 (1.01-1.16) | 0.021 | 0.11 |
| 2-year MACE-ALL | Low | 1.0 (ref) | n/a | n/a |
|  | Intermediate | 1.07 (1.01-1.13) | 0.017 | 0.10 |
|  | High | 1.06 (1.01-1.12) | 0.028 | 0.14 |
| 2-year coronary revascularization | Low | 1.0 (ref) | n/a | n/a |
|  | Intermediate | 1.02 (0.93-1.12) | 0.66 | 1.0 |
|  | High | 1.09 (0.99-1.19) | 0.071 | 0.43 |
| High-risk HEART scores (> 6, n = 3830) |  |  |  |  |
| 2-year MACE | Low | 1.0 (ref) | n/a | n/a |
|  | Intermediate | 0.99 (0.84-1.17) | 0.93 | 0.98 |
|  | High | 0.90 (0.76-1.06) | 0.22 | 0.88 |
| 2-year MACE-ALL | Low | 1.0 (ref) | n/a | n/a |
|  | Intermediate | 1.04 (0.90-1.21) | 0.58 | 1.0 |
|  | High | 0.98 (0.84-1.13) | 0.75 | 1.0 |
| 2-year coronary revascularization | Low | 1.0 (ref) | n/a | n/a |
|  | Intermediate | 1.05 (0.86-1.28) | 0.64 | 1.0 |
|  | High | 0.94 (0.77-1.15) | 0.55 | 1.0 |

Following stratification into HEART score risk categories (0 to 3 = low risk; 4 to 6 = moderate risk; > 6 = high risk), associations between outcomes and NICT referral group were assessed using mixed effects proportional hazard models adjusting for patient age, sex, risk factors (diabetes, coronary artery disease, coronary revascularization, smoking, hypertension,
hypercholesteremia, obesity, family history of premature coronary artery disease), peak troponin value while the emergency department, ischemic-appearing changes on electrocardiogram, and epoch, with censoring of competing outcomes (non-cardiac death for MACE, all-cause mortality for coronary revascularization), random effects at the facility level, and robust standard errors.
\* Holm-Bonferroni method for multiple comparisons within and between categories
Abbreviations: CV = cardiovascular; HEART = history, electrocardiogram, age, risk factors, troponin; MACE = major adverse cardiac event; MACE-ALL = major adverse cardiac event, including all-cause mortality; NICT = non-invasive cardiac test.

Sensitivity analyses excluding encounters from the third study epoch (Table S3), restricted to patients with quantifiable peak troponins (Table S4), or using mixed-effects logistic regression for the MACE-ALL outcome (Table S5) were consistent with the primary analysis. Post-hoc exploratory analysis restricted to the low and high NICT referral groups demonstrated statistically significant interactions between NICT referral group and age (p = 0.012), HEART risk score category (p = 0.032) and peak troponin level (p = 0.027), though in no instance was higher NICT referral significantly associated with a lower hazard of MACE (Figure 2). Unadjusted Kaplan-Meier curves for MACE-ALL are demonstrated in Figure 3.

**Figure 2.**
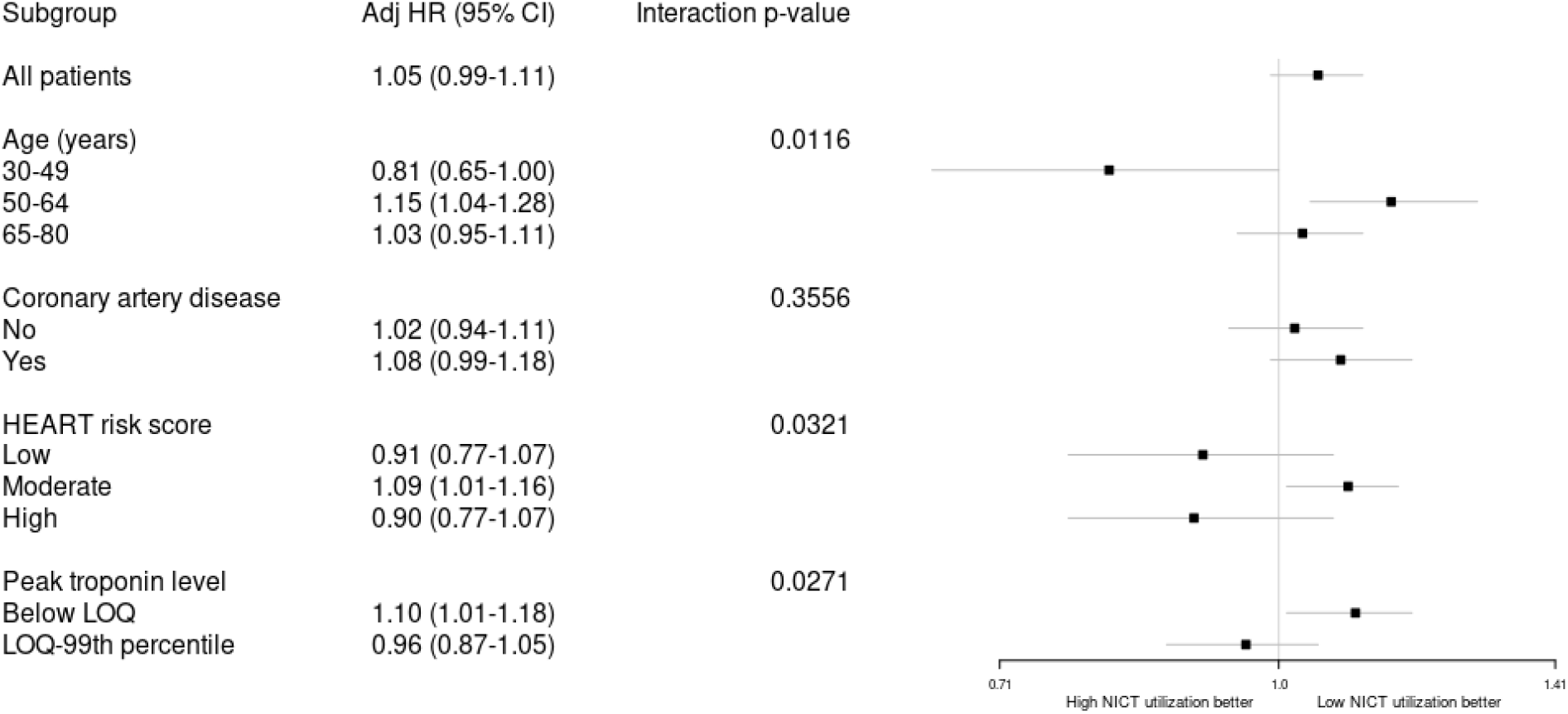
Forest plot of post-hoc exploratory interaction analyses for major adverse cardiac events (MACE): high versus low non-invasive cardiac test (NICT) referral group. Cause-specific mixed-effects proportional hazard models for MACE were run with an interaction term of NICT (high versus low) and each variable of interest (age, coronary artery disease, HEART risk score, peak troponin level) respectively, adjusting for other covariates with random effect at facility level and censoring for non-cardiac death. Abbreviations: Adj = adjusted; HEART = history, electrocardiogram, age, risk factors, troponin; LOQ = limit of quantitation; MACE = major adverse cardiac event; NICT = non-invasive cardiac test.

**Figure 3.**
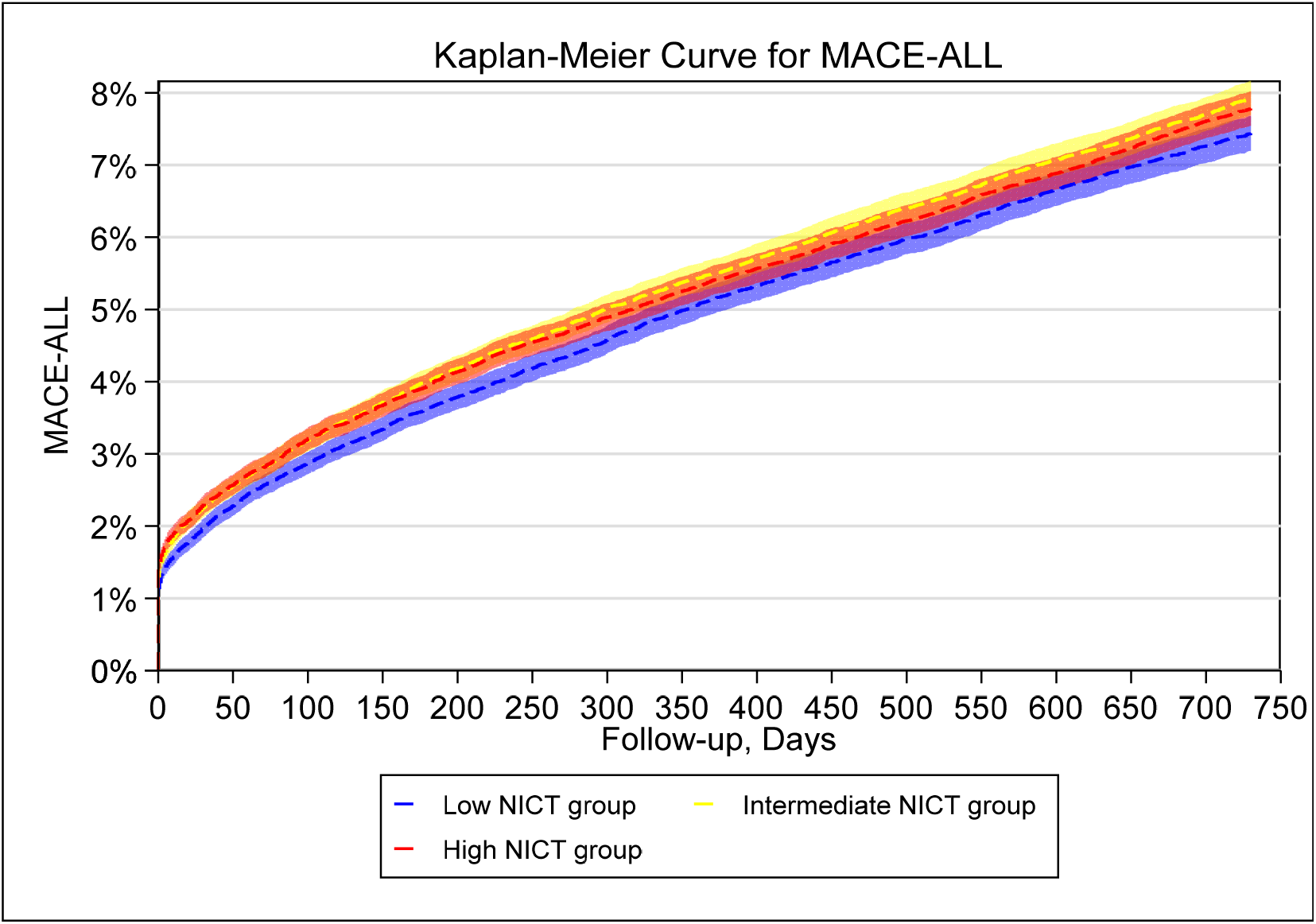
Kaplan-Meier curve. Unadjusted incidence of MACE-ALL within two years according to non-invasive cardiac test (NICT) referral group. MACE-ALL = major adverse cardiac event, inclusive of all-cause mortality; NICT = non-invasive cardiac test

## Discussion

The 2021 American Heart Association/American College of Cardiology chest pain evaluation guideline recommends against the urgent use of NICT in low-risk ED patients with chest pain, stating that “there is no evidence that stress testing or cardiac imaging within 30 days of the index ED visit improves their outcomes”.^38^ Since this recommendation was based on studies which used follow-up periods between 30 and 365 days ^4,6–8,10–12,27,39,40^, we sought to assess whether increased use of NICT might improve outcomes out to two years, specifically among ED patients without objective evidence of myocardial injury. Despite finding that 1 in 5 additional patients underwent objective cardiac testing within 30 days in the high NICT referral group (equating to approximately 10,000 additional patients, as compared to the low NICT referral group), we did not find that increased NICT referral was associated with a decrease in the hazard of two-year MACE or MACE-ALL. While the magnitude of effect of a covariate on an outcome cannot be directly inferred from proportional hazard model coefficients^37^, the lower bounded aHR 95% confidence intervals for MACE and MACE-ALL (0.99 to 1.02) make it unlikely that an inverse relationship exists between NICT referral and these outcomes, lending further support to the above recommendation.

The 2021 guideline also recommends further functional or anatomic cardiac testing for most patients at intermediate risk, defined as patients who do not meet low-risk criteria but have no evidence of acute myocardial injury by troponin testing, criteria that represent 52% of our study cohort. However, we did not observe any evidence that higher NICT referral reduced the hazard of MACE among moderate-risk patients (HEART score 4 to 6) despite similar between-group differences in 30-day objective cardiac testing and a higher incidence of two-year MACE (7.2% versus 4.8% in the whole cohort). While findings were also null for higher-risk patients (HEART score > 6 or quantifiable troponin measurements), both smaller sample sizes and less pronounced between-group differences in 30-day objective cardiac testing render those results more susceptible to a type 1 error, and thus less interpretable. We likewise did not observe any overall differences in coronary revascularizations with higher NICT referral, in contrast to our prior study of 60-day outcomes in this population but consistent with other longer-term studies of NICT.^16,17^ This may be attributable to a general decline in the performance of coronary revascularizations in the US over the past 20 years, as well as evidence suggesting equipoise between percutaneous coronary interventions and optimal medical therapy in patients with chronic coronary syndromes.^41–43^

While our findings differ from those of Roifmen et al.^5^ and Kawatakar et al.^7^, both of which reported some degree of reduction in MACE following NICT amongst high-risk ED patients with chest pain, there are some notable differences in study designs. Roifmen et al. matched on the propensity to have undergone NICT, rather than intent to refer for NICT, while Kawatkar et al. employed an instrumental variable approach (medical center-level variation in NICT performance and day-of-the week presentation). Both of these strategies are subject to residual confounding from factors associated with both NICT completion and outcomes, including access to care, health literacy, functional capacity, and treatment adherence, as well as MACE that preempt NICT completion.^44–46^ In contrast, by using *referral* for NICT as an indicator of practice variation at the individual physician level, the current study is more representative of an intention-to-treat design and avoids the potential for attrition bias when examining at the level of NICT and/or objective cardiac test completion.

To be clear, these findings do not indicate that patients without evidence of acute myocardial injury do not benefit from NICT, but rather that accepted determinants of clinical risk (such as the HEART score and its components) may not be ideally suited to identifying which patients might benefit. This is suggested by our post-hoc exploratory analysis where we observed an interaction between NICT group (low or high) and age, with patients aged 30 to 49 years bordering on a reduced hazard of MACE with high NICT referral (aHR 0.81, 95% CI 0.65-1.00). On the surface, this appears counterintuitive since this age group is largely comprised of low-risk patients (median age was 49 years in low-risk HEART score subgroup versus 65 years in the moderate-risk subgroup). However, it is also apparent that traditional cardiac risk factors carry greater prognostic weight in younger patients such that effective risk factor modification (such as smoking cessation), perhaps spurred on by greater referral for NICT, could have greater proportional benefits in younger individuals.^47–50^ But, given the low overall event rates among younger patients, well targeted approaches to identifying patients with occult and/or premature coronary artery disease are needed.^51^

The mode of NICT employed may also be an important determinant of potential benefit. Of the NICTs performed in our study, approximately one-third had an imaging component (myocardial perfusion, stress echocardiography or CT coronary angiography). The SCOT-HEART study, an open-label randomized controlled trial of 4146 patients with stable chest pain, compared CT coronary angiography to usual care and found significantly lower rates of mortality and myocardial infarction beginning at around 2 years among those randomized to the intervention.^40,52^ The CATCH trial, a randomized controlled trial of 576 patients with acute-onset chest pain and both normal ECGs and serial cardiac troponins, compared a CT coronary angiography-guided treatment strategy to standard care with a functional NICT and found that CT coronary angiography reduced downstream MACE out to a median of 19 months, though the difference in myocardial infarction or cardiac death was not statistically significant (p = 0.06).^53^ In contrast, the PROMSIE trial, which randomized 10,003 symptomatic outpatients without established coronary artery disease to either anatomic testing with CT coronary angiography or functional NICT, did not demonstrate any difference in clinical outcomes out to a median of two years.^54^ Whether patient selection for (and potential benefit from) CT coronary angiography can be further tailored with the use of high-sensitivity cardiac troponin assays (specifically among patients with mildly elevated but non-diagnostic high-sensitivity troponin values) is being addressed through the ongoing TARGET-CTCA study (NCT03952351). Thus, while the utility of routine and/or urgent functional NICT in low- and intermediate-risk ED patients has rightly been questioned, in large part due to low testing yields^55,56^, there may still yet be a role for anatomic NICT in mitigating long-term risks in selected patients.

This study has several limitations. All included patients were insured and cared for within an integrated health care system, which may have reduced overall outcome risks and somewhat limits generalizability. Given the observational study design, we cannot infer a causal relationship between NICT referral and outcomes, especially regarding any suggestion of an increased hazard of MACE and MACE-ALL with higher NICT referral, though it is plausible that higher NICT referral was associated with delayed diagnoses and treatment of alternative life-threatening conditions owing to anchoring bias.^57^ Though residual baseline confounding is possible, there were no observed imbalances in measured patient-level variables typically associated with NICT referral (e.g. age, risk factors, HEART score) and all study patients had active health plan insurance coverage and were grouped at the medical-center level over discrete epochs of time, making systematic differences in access to NICT between groups highly unlikely. Since our study excluded patients with troponin measurements above the 99^th^ percentile of normal, our results should not be extrapolated to this specific higher risk patient population in whom benefits of NICT are more likely.^7^ Finally, in terms of study power, though no formal calculations were performed given the fixed study period and uncertainty surrounding the ranges of NICT referral variation, the largest statistically significant result in terms of adjusted hazards (aHR 1.08 for MACE in the intermediate versus low NICT referral group, adjusted p value = 0.024) correlated to a 0.4% unadjusted absolute increase in two-year MACE between these groups. This finding suggests that the analysis was highly sensitive to clinically relevant differences in outcomes, with the caveat for smaller high-risk subgroups noted above.

## Conclusion

In this large observational study evaluating the association between 72-hour NICT referral and two-year outcomes following ED chest pain encounters in which acute myocardial injury had been excluded, higher NICT referral was not associated with a decreased risk of MACE. These findings suggest that routine NICT referral does not improve cardiac outcomes for most ED patients without evidence of acute myocardial injury, though the impact among the higher risk subset of patients remains unclear. Further studies in more targeted populations (using high-sensitivity troponin values) and/or using specific testing modalities (e.g. CT coronary angiography) may further inform best practices.

## Data Availability

Because of the sensitive nature of the data collected for this study, requests to access the dataset from qualified researchers trained in human subject confidentiality protocols may be sent to KPNC at.

## Non-standard Abbreviations and Acronyms

BMI: body mass index
CAD: coronary artery disease
ECG: electrocardiogram
ED: emergency department
HEART: history, electrocardiogram, age, risk factors, troponin
KPNC: Kaiser Permanente Northern California
LOQ: limit of quantitation
MACE: major adverse cardiac event
MACE-ALL: major adverse cardiac event, including all-cause mortality
NICT: non-invasive cardiac test
STEMI: ST-elevated myocardial infarction.

## Acknowledgments

None

## Supplemental Material

Supplemental Methods, Tables S1-S5

